# Identifying risk individuals for heart failure diagnosis within two years in the adult population in southern Sweden using gender, age, multimorbidity level, and socioeconomic status

**DOI:** 10.64898/2025.12.29.25343158

**Authors:** Mia Scholten, Anders Halling

## Abstract

**Objective:** Heart failure (HF) is a common disease among elderly individuals and is associated with poor quality of life and prognosis. Individuals at risk of developing HF are often already patients in primary healthcare, but it is often a difficult diagnosis at an early stage. Identifying patients at high risk for HF and initiating early treatment is crucial for their outcomes. Using the variables gender, age, multimorbidity (MM) level, and socioeconomic status (SES), we aimed to study the possibility of identifying individuals at high risk for HF diagnosis within two years in the adult population in southern Sweden.

**Design:** A register-based, cross-sectional cohort study.

**Setting and subjects:** 961,190 inhabitants aged 20 years and older without a HF diagnosis living in southern Sweden (Region Skåne) in 2015.

**Main outcome measure:** Predicting the risk for HF diagnosis within two years in Southern Sweden using the variables gender, age, MM level, and SES. SES was measured as CNI (Care Need Index) percentiles depending on the individuals listed at the primary healthcare centre.

**Results:** Age, MM level, and SES were added to the logistic model along with gender in steps. Each model was compared with the previous model using a likelihood-ratio test, which were all significant, resulting in an increased AUC (area under the curve) from 0.5144 to 0.9379. Age was the most important factor to predict the probability of HF diagnosis within two years. The second most important factor in our study was MM level. Both gender and SES improved the model significantly.

**Conclusions:** Age was the most important factor to predict probability of HF diagnosis within two years. The second most important factor in our study was MM level. Both gender and SES improved the model and its AUC significantly. Using the combination of these four variables was shown to be a possible tool to identify elderly individuals at high risk for HF diagnosis with high positive and negative predictive values in the adult population aged 70 years and older.

## Introduction

Heart failure (HF) is a common disease associated with age and constitutes a high healthcare burden globally. Most patients at risk of developing HF are already patients in primary healthcare, but the diagnosis could be difficult at an early stage. To identify the individuals at high risk for HF at an earlier stage could provide a shortcut to prevent complications and costs of this fatal disease, which also could improve the individuals’ quality of life. Patients with acute HF are reported to have a poor prognosis, comparable with those experiencing acute myocardial infarction [1]. Patients with stable chronic HF without treatment are also at high risk for fatal complications such as sudden death. Any delay of HF treatment initiation is associated with a worse outcome, and is therefore regarded as an important modifiable risk factor, because it is beneficial regardless of severity or duration of HF [1]. In summary, time to diagnosis and treatment plays a crucial role throughout the entire journey in HF patients, and we have an urgent need to better identify HF patients and provide adequate treatment in time. Current HF guidelines have also highlighted the importance of early diagnosis and treatment for HF patients to reduce disease progression and improve prognosis.

Many risk factors have been recognized for HF, most of which are associated with advancing age and MM level. Systemic aging has been acknowledged to influence various physiological processes contributing to structural and functional decline in cardiac tissue [2, 3]. Metabolic aging propelling intricate shifts in lipid and glucose metabolism, leading to lipid accumulation, insulin resistance, and mitochondrial dysfunction within cardiomyocytes, contributing to an increased incidence of left atrial dilation, atrial fibrillation, myocardial fibrosis, left ventricular hypertrophy, diastolic dysfunction, and elevated susceptibility to chronic HF in the elderly population [4, 5]. The age-related comorbidities like coronary artery disease (CAD), atherosclerosis, obesity, hypertension, diabetes, and chronic kidney disease (CKD) could further exacerbate HF [6]. CAD and atherosclerosis cause myocardial ischemia and dysfunction, while CKD-related fluid overload and uremic toxins aggravate HF through systemic inflammation and neurohormonal renin-angiotensin-aldosterone system (RAAS) activation [7, 8]. Hypertension drives cardiac hypertrophy and fibrosis [9]. Obesity-associated insulin resistance, hyperlipidemia, and inflammation contribute to cardiac dysfunction through metabolic disturbances [10]. The pathogenesis of the increased HF in elderly individuals could also partly be explained by long-term exposure to genomic, oxidative, epigenetic, autophagic, inflammatory, and regenerative stresses [11]. The accumulation of senescent cells with age implicates systemic low-grade chronic inflammation, aggravating the cardiac dysfunction, as well by promoting immune cell infiltration into the myocardium [12]. Even age-related changes of arterial stiffness, endothelial dysfunction, and impaired angiogenesis have an impact on heart dysfunction [13, 14]. Dysregulated nitric oxide signaling and mechanosignaling are also hallmarks for age-related vascular dysfunction contributing to HF [15, 16].

Since HF is strongly associated with MM level, we believe that the incidence of HF and its common comorbidities have a mutual effect on each other [17]. For example, the patients with the combination HF and type 2 diabetes (DM) were reported to have 37% (95% CI 35.9 - 38.1%) incidence of a third condition within five years, meanwhile HF patients with CKD as comorbidity only had 8.7% (95% CI 8.4 - 9.0%) incidence of a third condition within five years, most likely due to their high mortality rate (51.6%) [18]. Both CKD and DM are associated with an increased incidence of HF [19].

In this study, we aimed to facilitate the identification of individuals at high risk for developing HF over a two-year period using easily available information on risk factors for HF.

## Methods

### Data source and measurements

The Region Skåne healthcare register in southern Sweden provided data that contains anonymized registry information from the study population, including gender, age, diagnostic data, and SES. Data were collected concerning diagnoses at each consultation in all primary healthcare centres (PHCs) and secondary healthcare. Diagnoses were recorded according to the International Statistical Classification of Diseases and Related Health Problems version 10 (ICD 10) for a period of 18 months prior to the last week in 2015. HF was identified if the diagnosis code I50 was recorded, including all subtypes of HF.

### Multimorbidity

Individuals with at least two chronic conditions were considered to have multimorbidity. The study population was divided into 10 different MM levels: i.e., people with three chronic conditions belonged to MM level three; five chronic conditions belonged to MM level five; 10 or more chronic conditions belonged to MM level 10. A method developed by A Calderòn-Larrañaga *et al.* at the Aging Research Centre in Stockholm was used to identify chronic conditions [20]. They analysed the full list of ICD-10 codes on a four-digit level to define whether a diagnosis is chronic or not in an elderly population. To determine if a condition was chronic or not, the following key features were identified and discussed concerning their pertinence and sustainability in older populations: duration, course, reversibility, treatment, and consequences. They were then grouped into 60 groups of chronic conditions if their duration exceeded three months. We applied their definition and list of chronic conditions to estimate the MM level in our study population. All information about diagnoses was obtained from electronic medical record database in the county council in Scania. MM level was then estimated by counting the number of chronic conditions in each patient and divided into ten groups. The groups with two chronic conditions or more were considered multimorbid, and group 10 had at least 10 chronic conditions.

### Socioeconomic status

SES was measured as Care Need index (CNI) [21] to divide the PHCs at which the individuals were listed into 10 groups depending on their CNI. CNI is based on different measures of a group, in this case the patients listed at the PHCs in Region Skåne. Most people are listed at the PHCs in their neighbourhood, which has an average CNI level. CNI 1 was assigned to those patients listed at PHCs who belonged to the most socioeconomically affluent percentile, and CNI 10 was assigned to those patients listed at PHCs who belonged to the most socioeconomically deprived percentile [21].

### Statistical analyses

We analyzed the population living in Region Skåne during 2015 (981, 383), of whom only those aged 20 years and older without HF diagnosis were included (961,190). From a total of 961,190 study participants living in Region Skåne at the end of 2015 (31^st^ December 2015), 50.9% were women and 49.1% were men. The variables we used to characterize this study population were gender, age, MM level, and SES (CNI).

We used frequencies, percentages, and cross tabulations for descriptive analysis. Logistic regression was used to analyse the OR for a HF diagnosis within two years using multivariate models. A p-value of < 0.05 was considered statistically significant.

The primary outcome was to identify individuals at high risk for HF diagnosis within two years, which was analyzed using logistic regression. Linear predictions were made based on models by adding variables in steps (Model 1 – Model 4), e.g., gender, age, MM level, and SES. Each model was compared with the previous model using a likelihood-ratio test, which were all significant. ROC (Receiver Operating Characteristic) and AUC were used to compare different models’ predictive value of the four variables for probability of HF diagnosis [22]. A heatmap was used to illustrate the predicted percentage probabilities according to age and MM level. To find the optimal cut-off point to measure sensitivity and specificity, the Youden method was used. Positive and negative predictive values were calculated using the prevalence in the whole study population and for those aged 70 and older.

We used STATA version 19.5 (Stata Corporation, Texas, USA) for statistical analyses.

### Statement of ethics

Data in the present study are based on anonymized information provided by Region Skåne. They provided anonymized information for research purposes once the study had been approved by the Ethics Committee at Lund University (application no. 2023-03587-01). All analyses were performed in accordance with relevant guidelines and regulations. Due to the requirement for anonymized data, each individual could not be asked for consent to participate; active refusal of participation was instead applied. This was done by publishing information about the planned study in the Swedish local newspaper “Sydsvenskan”. The advertisement outlined the study and contained information on how to contact the research manager (first author) to opt out of the study.

## Results

The proportions of diagnosed HF within two years according to age, gender, MM level, and CNI are presented in Table 1. The most important variable was age and the second most important was MM level (Table 2). 84.4% of those who developed HF had multimorbidity two years earlier. The OR for receiving a HF diagnosis within two years rose sharply with advancing age and MM level. Men had a higher OR for HF than women. Among all the CNI levels, the highest OR for HF was seen in the most socioeconomically deprived group.

**Table 1.**
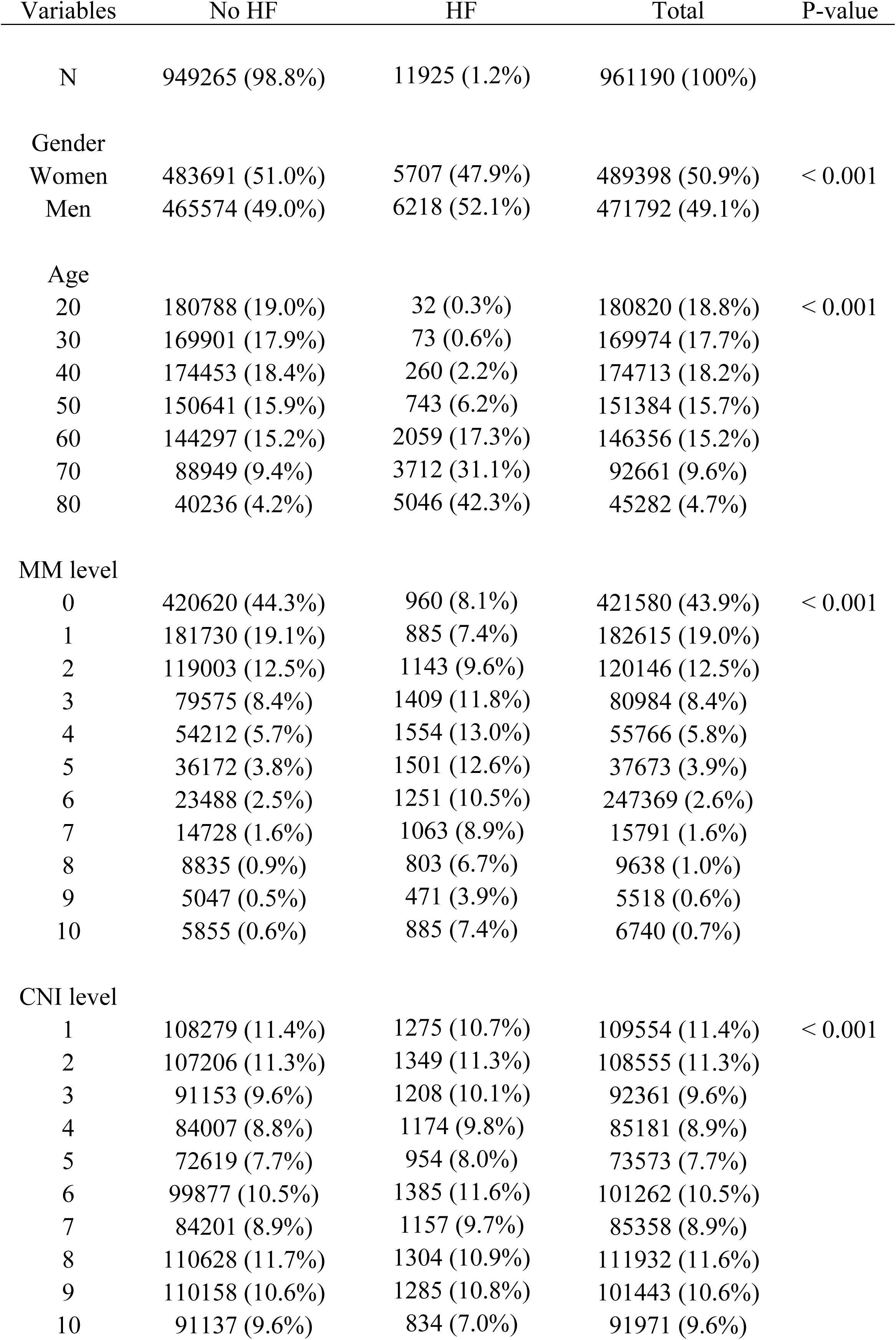
Proportions of diagnosed heart failure in Region Skåne within two years, categorized by gender, age, multimorbidity levels, and socioeconomic status (CNI levels).

**Table 2.** Odds ratios for heart failure diagnosis within two years, depending on different variables in the adult population in Region Skåne.

| Variables | OR (HF) | 95% CI | P - value |
| --- | --- | --- | --- |
| Gender |  |  |  |
| Women | 1 |  |  |
| Men | 1.55 | 1.49 - 1.61 | < 0.01 |
| age |  |  |  |
| 20 | 1 |  |  |
| 30 | 2.32 | 1.53 - 3.51 | < 0.01 |
| 40 | 7.51 | 5.20 - 10.84 | < 0.01 |
| 50 | 21.11 | 14.80 - 30.09 | < 0.01 |
| 60 | 50.13 | 35.30 - 71.20 | < 0.01 |
| 70 | 117.38 | 82.68 - 166.64 | < 0.01 |
| 80 | 332.60 | 227.22 - 458.01 | < 0.01 |
| MM level |  |  |  |
| 0 | 1 |  |  |
| 1 | 1.26 | 1.15 - 1.38 | < 0.01 |
| 2 | 1.65 | 1.51 - 1.80 | < 0.01 |
| 3 | 2.11 | 1.94 - 2.30 | < 0.01 |
| 4 | 2.64 | 2.43 - 2.88 | < 0.01 |
| 5 | 3.21 | 2.94 - 3.49 | < 0.01 |
| 6 | 3.58 | 3.28 - 3.92 | < 0.01 |
| 7 | 4.41 | 4.02 - 4.85 | < 0.01 |
| 8 | 5.21 | 4.71 - 5.77 | < 0.01 |
| 9 | 5.16 | 4.58 - 5.81 | < 0.01 |
| 10 | 7.98 | 7.21 - 8.83 | < 0.01 |
| CNI level |  |  |  |
| 1 | 1 |  |  |
| 2 | 1.07 | 0.99 - 1.16 | > 0.05 |
| 3 | 1.16 | 1.07 - 1.26 | < 0.01 |
| 4 | 1.20 | 1.10 - 1.30 | < 0.01 |
| 5 | 1.23 | 1.13 - 1.34 | < 0.01 |
| 6 | 1.15 | 1.07 - 1.25 | < 0.01 |
| 7 | 1.24 | 1.14 - 1.34 | < 0.01 |
| 8 | 1.18 | 1.09 - 1.28 | < 0.01 |
| 9 | 1.36 | 1.25 - 1.47 | < 0.01 |
| 10 | 1.40 | 1.28 - 1.54 | < 0.01 |

OR = odds ratio

HF = heart failure, CI = confidence interval, MM = multimorbidity, CNI = Care Need Index OR = odds ratio

Age, MM level, and SES were added to the logistic model with gender in steps. Each model was compared with the previous model using a log likelihood test and was seen to increase AUC significantly from 0.51 to 0.94 (Fig. 1). AUC for gender (Model 1) was 0.5144, for gender and age (Model 2), it was 0.9213, and for gender, age, and MM level (Model 3), it was 0.9376. For the combination of all four variables, gender, age, MM level, and SES (Model 4), the AUC was 0.9379 (Fig. 1). Age was the most important factor to predict the probability of HF diagnosis within two years. The second most important factor in our study was MM level. Gender and SES also added to the model significantly.

**Fig. 1.**
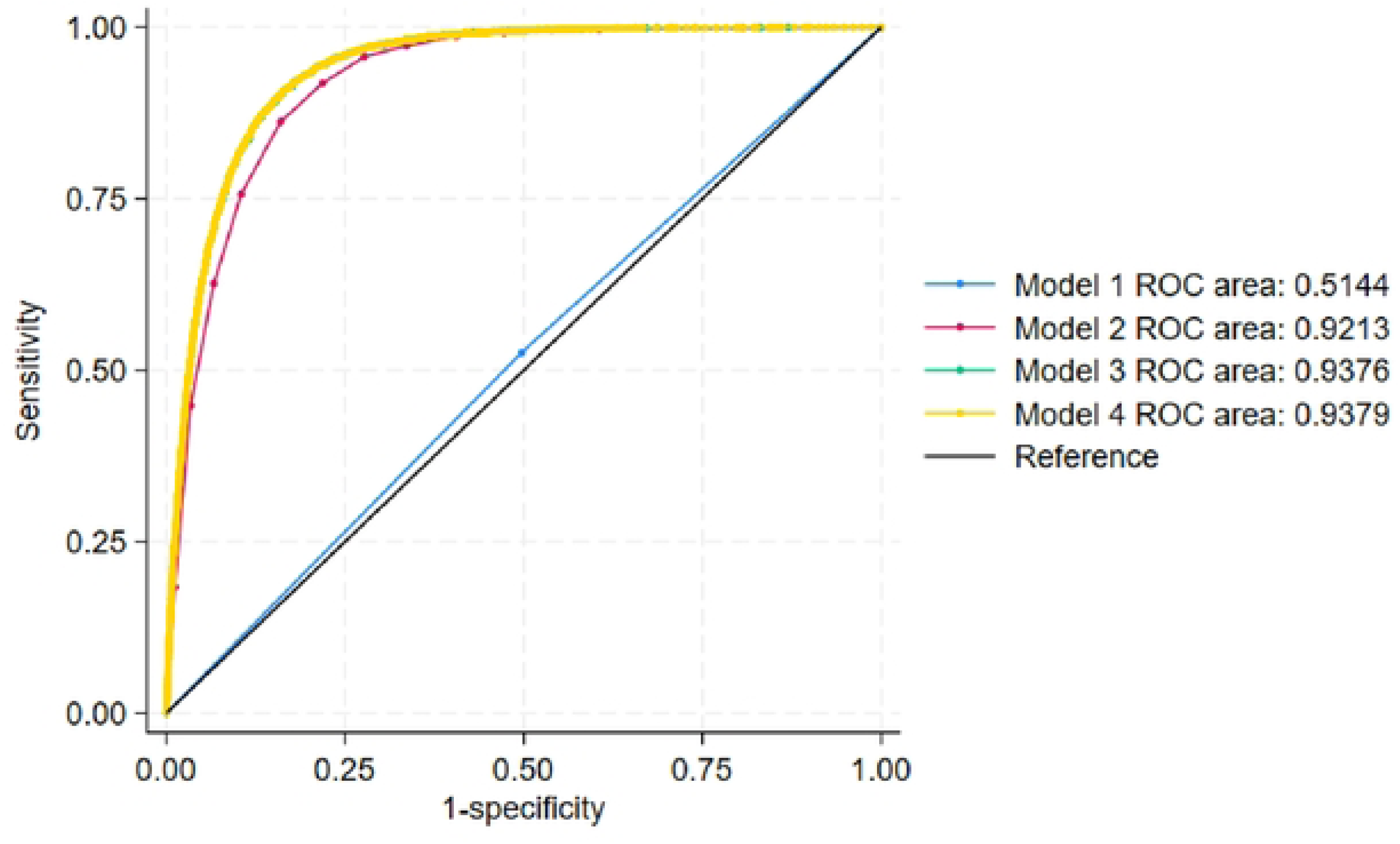
ROC (Receiver Operating Characteristic) curves comparing four models of different complexity, Models 1 - 4. Model 1 = gender Model 2 = gender and age Model 3 = gender, age, and multimorbidity level Model 4 = gender, age, multimorbidity level, and socioeconomic status (CNI level) The heatmap in Fig. 2 showed that the predicted percentage probabilities (Model 4) were approximately highest at the combination of age over 70 and MM level five or higher.

**Fig. 2.**
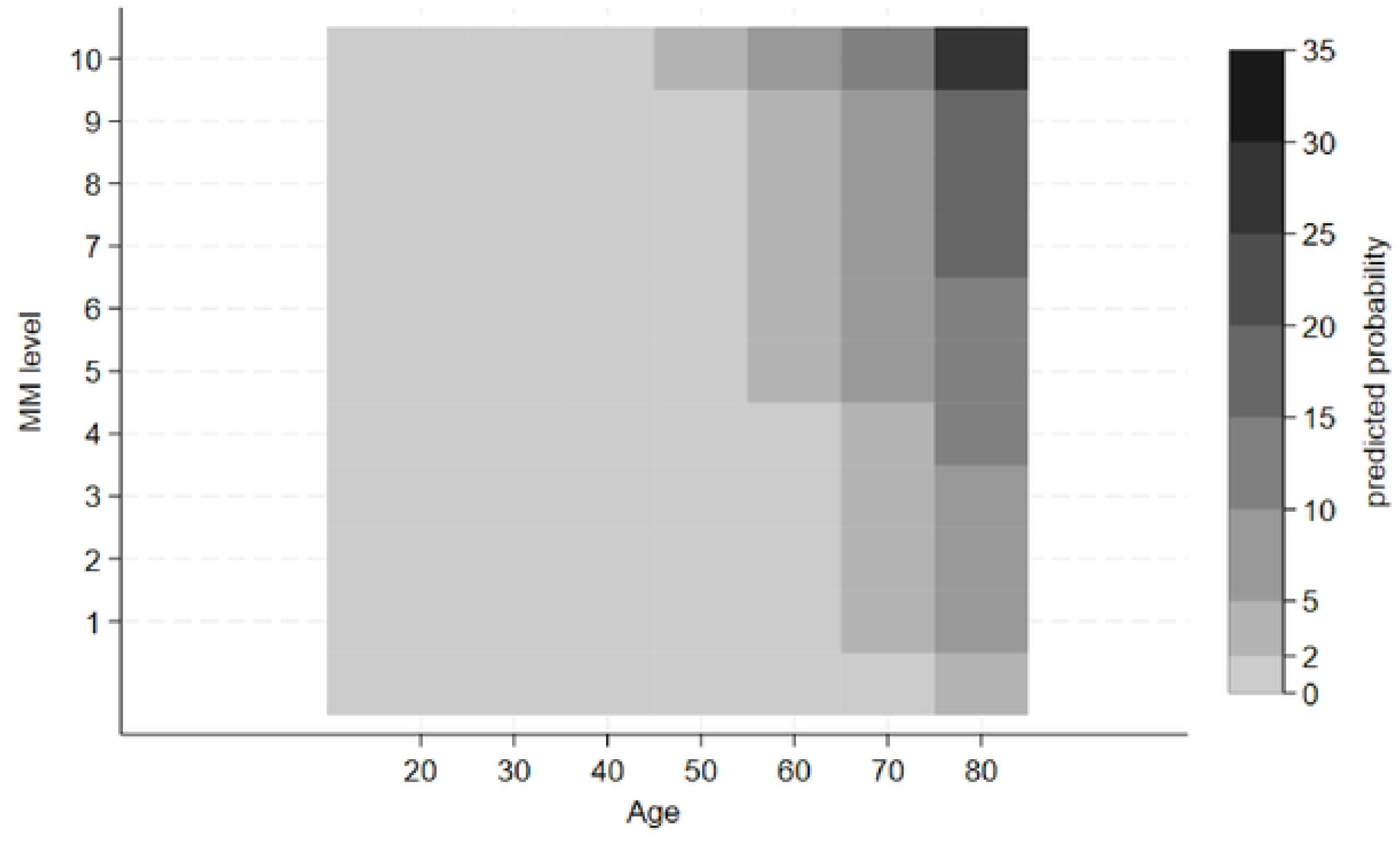
Heatmap of the predicted percent probabilities (Model 4) for heart failure diagnosis according to multimorbidity levels. When all four variables were in the model, an optimal cutoff point according to Youden was established at 1.15 percent predicted probability with a sensitivity of 87.69% and specificity of 78.48%. The positive- and negative predictive values for the whole adult population was 4.78% and 99.81%; for those aged 70 years and older, it was 21.02% and 98.99%; and for those aged 80 years and older, it was 33.62% and 98.09%, respectively.

## Discussion

Age was found to be the most important factor to predict a two-year risk of HF diagnosis for an individual. The second most important factor in our study was MM level. Gender and SES also added the predictive value significantly to this model. AUC for gender was only 0.5144 when calculated with ROC, but increased to 0.9213 when age was added as a variable. The area under the ROC curve increased further to 0.9376 when the combination of gender, age and, MM level was calculated together. Ultimately, when all four variables were included in the ROC, the AUC reached 0.9379. The positive- and negative predictive values for the whole adult population was 4.78% and 99.81; for those aged 70 years and older, it was 21.02% and 98.99%; and for those aged 80 years and older, it was 33.62% and 98.09%, respectively.

A wide variety of comorbidities are expected to contribute differently to the HF diagnosis, which have more impact than SES and gender, but less than advancing age. Interestingly, the difference in OR for HF diagnosis was obvious between the genders after adjusting for MM, age, and SES (CNI), which is in line with our published results from this study population: the elderly socioeconomically deprived men with MM were more prone to develop HF. Ischemic heart disease, which is recognized as the most common etiology of HF in Western countries, affects men to a greater extent and consequently contributes to the higher OR for HF diagnosis in men than women [23, 24]. The most socioeconomically deprived group in Region Skåne had the highest probability for HF and only 33% were over 50 years old, thus indicating that these individuals have more potential to improve their outcomes than the more socioeconomically affluent population, when hypothesizing that advancing age is the strongest risk factor for HF [17].

The very pronounced increase in OR for HF diagnosis between age group 70 and 80 is most likely the result of a synergetic effect of MM level, socioeconomic deprivation, and the age-related chronic HF process, like inflammation, mitochondrial dysfunction, senescence, and declining cardiomyocyte regeneration [25]. The summary effect of MM level and socioeconomic deprivation may affect the biological age of the individual. The complexity of different comorbidities supposedly causes an elevated susceptibility to chronic HF as well.

Our previous cross-sectional study on this population revealed the probability for HF increased substantially with advancing age and MM level, which certainly explains the highest OR for HF diagnosis within two years in the age group 80. These data could be interpreted as HF diagnosis is associated with advancing age and mortality rate, presumably as consequences of MM. A British observational study reported that 16% of HF patients had DM and CKD, who had at least three times higher mortality rates (78.6%) compared with patients with HF only (20.5%), HF with CKD (24.9%), or diabetes (20.5%) as comorbidity [26].

A cohort study including 24,675 participants without HF diagnosis, stratified by age into young (< 55 years; n = 11,599), middle-aged (55 - 64 years; n = 5,587), old (65 - 74 years; n = 5,190), and elderly (≥ 75 years; n = 2,299) individuals [27]. Over the median follow-up time of 12.7 years, 1% of young, 5% of middle-aged, 10% of old, and 18% of elderly participants developed HF. 32% (n = 44) of the HF cases in young participants were classified as HFpEF compared with 43% (n = 179) in elderly participants [27]. Risk factors, including diabetes, hypertension, current smoking history, and previous myocardial infarction, implicated a greater relative risk in younger compared with older participants (P for interaction < 0.05 for all). For example, hypertension caused more than double the risk of HF development in young participants (HR 3.02, 95% CI 2.10 - 4.34; P < 0.001) than in elderly participants (HR 1.43, 95% CI 1.13 - 1.81; P = 0.003) [27]. These younger participants had a lower MM burden compared to the elderly, which could explain their lower incidence of HF and a higher relative risk for HF development of each comorbidity. The absolute risk for developing HF was lower in younger than in older participants with and without risk factors, which could be explained by the age-related changes in the myocardium in elderly individuals fostering HF development [27].

Recognizing biological aging as a major modifiable risk factor for HF has guided us to target systemic aging through both lifestyle changes and medical therapy with enhanced attention on the individuals at high risk for HF. Primary healthcare could contact these people to initiate treatment and prevent HF or its complications. Lifestyle interventions like exercise with acknowledged antioxidant effects can reduce mitochondrial dysfunction, senescence, and inflammation, hence stimulate cardiomyocyte regeneration and partly restore the pathological cardiac remodeling in age-related chronic HF [28, 29]. Adherence to plant-based and ketogenic diets is also associated with lower HF risk [30, 31]. Caloric restriction leading to reduced adiposity improves myocardial function by reducing body fat, which is a pioneering therapy for overweight HF patients [32]. Medical therapy, including senolytics and senomorphics to reduce senescence, antioxidants targeting mitochondrial stress, anti-inflammatory drugs like interleukin (IL)-1β inhibitors, metabolic regulators such as nicotinamide riboside, resveratrol and sirtuin (SIRT) activators and autophagy enhancers like metformin and sodium-glucose cotransporter 2 (SGLT2) inhibitors, are all available treatment options for preserving cardiac function and mitigating the age-related HF burden [25].

## Strengths and limitations

The strongest strength is the similar results compared with the calculations based on the same study population from our previous study. Most P-values had a high level of statistical significance, which could be interpreted with substantial accuracy. Area under the ROC curve was high (0.94), although only four easily available variables were analyzed in the logistic regression, which facilitates the future project to identify individuals at risk for HF. These results could guide us to differentiate and prioritize between many risk factors on the way to identifying individuals at high risk for HF.

This study has limitations. The combination of different comorbidities could be more clearly defined in order to facilitate the identification of typical HF patients. For example, CAD, hypertension, chronic obstructive pulmonary disease (COPD), atrial fibrillation, CKD and obesity probably have more impact on HF development than other comorbidities, like cataract, arthritis and dementia. The severity of the comorbidities could also have different impact on the risk for HF. Our calculations did not consider the continuous increase of the population living in Region Skåne. Another error source could be the dynamic situation in CNI classification during the study period, because all patients can be listed at PHCs as a choice of their own and not always at the place of their neighbourhood. Patients with MM usually undergo medical treatments, but the quality was not evaluated, which could influence our results. Those risk factors for HF like obesity or hypertension, which do not cause immediate disability, could easily be neglected by the patients, thus resulting in delayed treatment.

## Conclusion

Aging was the major risk factor for HF diagnosis within two years, followed by MM, gender, and SES in our study. Using the combination of these four variables in a population sample was found to be a reliable tool to identify individuals at high risk for HF diagnosis.

## Data Availability

All relevant data are within the manuscript and its Supporting Information files.

## Acknowledgements

We thank Region Skåne for providing the patient data to enable this study. We are indebted to Patrick O’Reilly for his expertise and invaluable advice in proofreading the manuscript.

## Consent for publication

Not applicable.

## Statement of contribution

In accordance with the Vancouver Protocol, AH was involved in data collection, design of the study, data analysis and editing the manuscript. MS contributed with data analysis, writing, and editing the manuscript.

## References

1. Abdin A, Anker SD, Butler J, Coats AJS, Kindermann I, Lainscak M, Lund LH, Metra M, Mullens W, Rosano G et al: ‘Time is prognosis’ in heart failure: time-to-treatment initiation as a modifiable risk factor. ESC Heart Fail 2021, 8(6):4444–4453.

2. Strait JB, Lakatta EG: Aging-associated cardiovascular changes and their relationship to heart failure. Heart Fail Clin 2012, 8(1):143–164.

3. Triposkiadis F, Xanthopoulos A, Butler J: Cardiovascular Aging and Heart Failure: JACC Review Topic of the Week. J Am Coll Cardiol 2019, 74(6):804–813.

4. Xie S, Xu SC, Deng W, Tang Q: Metabolic landscape in cardiac aging: insights into molecular biology and therapeutic implications. Signal Transduct Target Ther 2023, 8(1):114.

5. Bhashyam S, Parikh P, Bolukoglu H, Shannon AH, Porter JH, Shen YT, Shannon RP: Aging is associated with myocardial insulin resistance and mitochondrial dysfunction. Am J Physiol Heart Circ Physiol 2007, 293(5):H3063–3071.

6. Screever EM, van der Wal MHL, van Veldhuisen DJ, Jaarsma T, Koops A, van Dijk KS, Warink-Riemersma J, Coster JE, Westenbrink BD, van der Meer P et al: Comorbidities complicating heart failure: changes over the last 15 years. Clin Res Cardiol 2023, 112(1):123–133.

7. Szlagor M, Dybiec J, Młynarska E, Rysz J, Franczyk B: Chronic Kidney Disease as a Comorbidity in Heart Failure. Int J Mol Sci 2023, 24(3).

8. Frostegård J: Immunity, atherosclerosis and cardiovascular disease. BMC Med 2013, 11:117.

9 . Forrester SJ, Booz GW, Sigmund CD, Coffman TM, Kawai T, Rizzo V, Scalia R, Eguchi S: Angiotensin II Signal Transduction: An Update on Mechanisms of Physiology and Pathophysiology. Physiol Rev 2018, 98(3):1627–1738.

10. Ebong IA, Goff DC, Jr., Rodriguez CJ, Chen H, Bertoni AG: Mechanisms of heart failure in obesity. Obes Res Clin Pract 2014, 8(6):e540–548.

11. Li H, Hastings MH, Rhee J, Trager LE, Roh JD, Rosenzweig A: Targeting Age-Related Pathways in Heart Failure. Circ Res 2020, 126(4):533–551.

12. Franceschi C, Garagnani P, Parini P, Giuliani C, Santoro A: Inflammaging: a new immune-metabolic viewpoint for age-related diseases. Nat Rev Endocrinol 2018, 14(10):576–590.

13. Ya J, Bayraktutan U: Vascular Ageing: Mechanisms, Risk Factors, and Treatment Strategies. Int J Mol Sci 2023, 24(14).

14. Cornuault L, Rouault P, Duplàa C, Couffinhal T, Renault MA: Endothelial Dysfunction in Heart Failure With Preserved Ejection Fraction: What are the Experimental Proofs? Front Physiol 2022, 13:906272.

15. Wang L, Luo JY, Li B, Tian XY, Chen LJ, Huang Y, Liu J, Deng D, Lau CW, Wan S et al: Integrin-YAP/TAZ-JNK cascade mediates atheroprotective effect of unidirectional shear flow. Nature 2016, 540(7634):579–582.

16. Layland J, Li JM, Shah AM: Role of cyclic GMP-dependent protein kinase in the contractile response to exogenous nitric oxide in rat cardiac myocytes. J Physiol 2002, 540(Pt 2):457–467.

17. Scholten M, Midlöv P, Halling A: Disparities in prevalence of heart failure according to age, multimorbidity level and socioeconomic status in southern Sweden: a cross-sectional study. BMJ Open 2022, 12(3):e051997.

18. Pasea L, Dashtban A, Mizani M, Bhuva A, Morris T, Mamza JB, Banerjee A: Risk factors, outcomes and healthcare utilisation in individuals with multimorbidity including heart failure, chronic kidney disease and type 2 diabetes mellitus: a national electronic health record study. Open Heart 2023, 10(2).

19. Perkovic V, Agarwal R, Fioretto P, Hemmelgarn BR, Levin A, Thomas MC, Wanner C, Kasiske BL, Wheeler DC, Groop PH: Management of patients with diabetes and CKD: conclusions from a “Kidney Disease: Improving Global Outcomes” (KDIGO) Controversies Conference. Kidney Int 2016, 90(6):1175–1183.

20. Calderon-Larranaga A, Vetrano DL, Onder G, Gimeno-Feliu LA, Coscollar-Santaliestra C, Carfi A, Pisciotta MS, Angleman S, Melis RJF, Santoni G et al: Assessing and Measuring Chronic Multimorbidity in the Older Population: A Proposal for Its Operationalization. J Gerontol A Biol Sci Med Sci 2017, 72(10):1417–1423.

21. Sundquist K, Malmström M, Johansson S-E, Sundquist J: Care Need Index, a useful tool for the distribution of primary health care resources. Journal of Epidemiology and Community Health 2003, 57(5):347–352.

22. Andrich D: Georg Rasch and Benjamin Wright’s Struggle With the Unidimensional Polytomous Model With Sufficient Statistics. Educ Psychol Meas 2016, 76(5):713–723.

23. Khatibzadeh S, Farzadfar F, Oliver J, Ezzati M, Moran A: Worldwide risk factors for heart failure: a systematic review and pooled analysis. Int J Cardiol 2013, 168(2):1186–1194.

24. Lam CSP, Arnott C, Beale AL, Chandramouli C, Hilfiker-Kleiner D, Kaye DM, Ky B, Santema BT, Sliwa K, Voors AA: Sex differences in heart failure. Eur Heart J 2019, 40(47):3859–3868c.

25. Fang Z, Raza U, Song J, Lu J, Yao S, Liu X, Zhang W, Li S: Systemic aging fuels heart failure: Molecular mechanisms and therapeutic avenues. ESC Heart Fail 2025, 12(2):1059–1080.

26. Lawson CA, Seidu S, Zaccardi F, McCann G, Kadam UT, Davies MJ, Lam CS, Heerspink HL, Khunti K: Outcome trends in people with heart failure, type 2 diabetes mellitus and chronic kidney disease in the UK over twenty years. EClinicalMedicine 2021, 32:100739.

27. Tromp J, Paniagua SMA, Lau ES, Allen NB, Blaha MJ, Gansevoort RT, Hillege HL, Lee DE, Levy D, Vasan RS et al: Age dependent associations of risk factors with heart failure: pooled population based cohort study. Bmj 2021, 372:n461.

28. Danduboyina A, Panjiyar BK, Borra SR, Panicker SS: Cardiovascular Benefits of Resistance Training in Patients With Heart Failure With Reduced Ejection Fraction: A Systematic Review. Cureus 2023, 15(10):e47813.

29. Isath A, Koziol KJ, Martinez MW, Garber CE, Martinez MN, Emery MS, Baggish AL, Naidu SS, Lavie CJ, Arena R et al: Exercise and cardiovascular health: A state-of-the-art review. Prog Cardiovasc Dis 2023, 79:44–52.

30. Lara KM, Levitan EB, Gutierrez OM, Shikany JM, Safford MM, Judd SE, Rosenson RS: Dietary Patterns and Incident Heart Failure in U.S. Adults Without Known Coronary Disease. J Am Coll Cardiol 2019, 73(16):2036–2045.

31. Montgomery RL, Potthoff MJ, Haberland M, Qi X, Matsuzaki S, Humphries KM, Richardson JA, Bassel-Duby R, Olson EN: Maintenance of cardiac energy metabolism by histone deacetylase 3 in mice. J Clin Invest 2008, 118(11):3588–3597.

32. Bianchi VE: Caloric restriction in heart failure: A systematic review. Clin Nutr ESPEN 2020, 38:50–60.

